# Curating genomic disease-gene relationships with Gene2Phenotype

**DOI:** 10.1101/2024.02.26.24303357

**Authors:** T Michael Yates, Morad Ansari, Louise Thompson, Sarah E Hunt, Elena Cibrian Uhalte, Rachel J Hobson, Joseph A Marsh, Caroline F Wright, Helen V Firth

## Abstract

Genetically determined disorders are highly heterogenous in clinical presentation and underlying molecular mechanism. The evidence underpinning these conditions in the peer-reviewed literature is variable and requires robust critical evaluation for diagnostic use. Here, we present a structured curation process for the Gene2Phenotype (G2P) project. This draws on multiple lines of clinical, bioinformatic and functional evidence. The process utilises and extends existing terminologies, allows for precise definition of the molecular basis of disease, and confidence levels to be attributed to a given gene-disease assertion. In-depth disease curation using this process will prove useful in applications including in diagnostics, research and the development of targeted therapeutics.

## Background

Genomic sequencing is widely used in the diagnosis of rare genetically determined disorders. It is relatively straightforward and cost-effective to generate large volumes of data from genome or exome sequencing. However, sampling of a large genomic footprint inevitably results in a number of candidate disease-associated variants passing initial filtering steps. It is therefore important to develop automated strategies to reduce false negative and false positive results, in order to optimise use of clinician and scientist time and focus attention on variants with the highest likelihood of being clinically significant. The Gene2Phenotype (G2P) database (1) was developed to enable high-throughput filtering of variant calls and prioritisation of likely clinically relevant variants (2). G2P has been successfully used in a number of diagnostic clinical and research applications, for example the DDD (Deciphering Developmental Disorders) Study (3), assessment of disorders of the eye (4) and inherited cardiac disorders (5).

G2P defines monogenic gene-disease associations through Locus-Genotype-Mechanism-Disease-Evidence (LGMDE) threads (2). This allows for precise definition of the clinical phenotype and molecular basis of a given condition. G2P was developed in 2012, primarily as a database of all known loci associated with Developmental Disorders (DDG2P). DDG2P has more than tripled in size over the last decade, now covering over 2500 loci (6). The system was designed to be generalizable across disease domains, and has now expanded to include Cancer, Cardiac, Eye, Skeletal and Skin disorders (Figure 1). Each panel is freely downloadable (1), comprehensive and actively curated by expert curators. G2P entries can be present in multiple panels for convenience.

**Figure 1.**
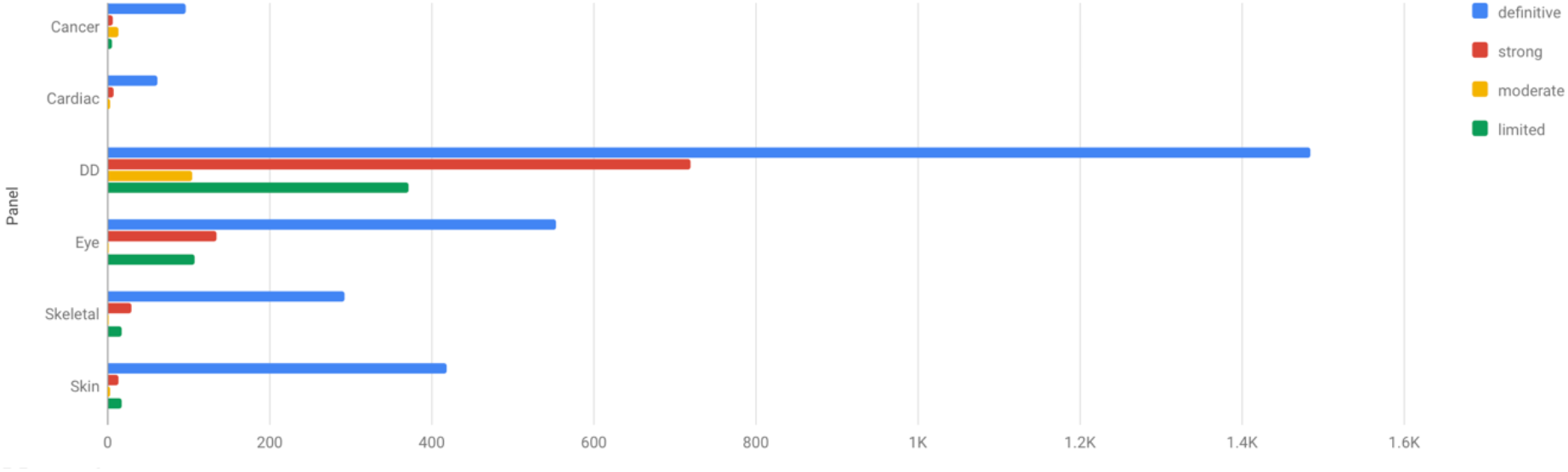
Gene disease pair counts for each confidence level per G2P panel (1). Downloaded 31^st^ January,2024.

Each G2P entry is curated by clinical and scientific experts through detailed assessment of publications from the peer-reviewed literature. For newly described gene-disease associations, manuscripts are identified through a monthly manual search of relevant journals. Case reports/case series containing detailed human phenotypic data are prioritised. This is an important process given the significant variation in evidence available for different gene-disease assertions. A curated confidence level for this assertion is assigned, to enable prioritisation of clinically relevant diagnostic variants.

## Methods

Here, we present a newly defined G2P curation process with associated data structures. These have been revised, to include granular molecular and mechanistic data to allow more precise disease definition, as well as to use standardised terminology compatible with the Gene Curation Coalition (GenCC) standards (7). A formalised curation template has been developed, which we present as a resource for the genomics community (Supplementary Materials). The template was iteratively developed by the DDG2P curation team, comprising clinical geneticists, clinical scientists, bioinformaticians and specialists in biomedical data curation. Input was then sought at a workshop where a wide range of expertise was represented, including academic experts in clinical genetics and genomics, biomedical informatics, computational protein biology, and genome annotation.

This curation process results in a focused, in-depth assessment of the evidence underlying the association of a disease with a particular genomic locus. Phenotypic and molecular data is analysed in a consistent, sequential manner following the curation template. This populates the LGMDE threads as well as allowing for a confidence assertion to be made.

## Results

### LGMDE threads

Each LGMDE thread is described here in detail, including definitions, terminology, data sources, and utilisation in the curation process.

### Locus

The locus thread is typically a gene or can be a genomic interval (defined as chromosome:genomic coordinates for a given reference genome assembly). For genes, the HUGO Gene Nomenclature Committee (HGNC) (8) symbol is used for ease of reference. The symbol is mapped to the relevant stable numerical HGNC ID.

### Genotype

#### Allelic requirement

Standardised Human Phenotype Ontology (HPO) (9) allelic requirement terms are used. These have corresponding Mendelian inheritance terms. For example, monoallelic_autosomal – Autosomal dominant – HP:0000006. In general, genes which have been associated with multiple allelic requirements for a given disease require separate G2P entries. This would apply, for example, to monoallelic_autosomal and biallelic_autosomal disorders. X-linked conditions which are usually penetrant in males and recessive in females may be recorded as monoallelic_X_hemizygous. X-linked disease where heterozygous females and hemizygous males have similar phenotypes, for example in relation to SHOX and *SMC1A* variants, are recorded as monoallelic_X_heterozygous. However, it is recognized that this distinction may be difficult in practice, and separate entries can be used after discussion by the curation group.

#### Cross-cutting modifier

Optional cross-cutting modifiers give extra information for a gene-disease relationship. The terms used largely correspond to children of “Inheritance qualifier” (HP:0034335), for example, “Typically de novo” (HP:0025352). A subset of these HPO terms is included in the curation template. These have been chosen to focus curation on collection of data most relevant to diagnostic filtering.

Two additional cross-cutting modifiers not representing inheritance information are defined. The first is for alerting the user to potential secondary findings (including American College of Medical Genetics Secondary Findings (10) and/or late onset conditions). The second modifier is for a “Restricted Variant Set”. This may include, for example, where diseases are associated with a single recurrent variant, or variants only found in a particular protein domain. Further information regarding the specific variant set is recorded elsewhere in the curation data.

#### Types of variants reported

A comprehensive list of Sequence Ontology (11,12) terms is used to record the types of variants reported in association with a disease entity. For example, “frameshift_variant” (SO:0001589). (11). Additional information is recorded for each variant type, including: if it is reported as *de novo* and/or inherited and/or of unknown inheritance, whether it is predicted to escape or trigger nonsense mediated decay (NMD) and gene domain/genomic region. This is important for filtering genomic data, as well as in defining the mechanism of disease.

#### Protein view

A snapshot of the DECIPHER (13) protein view for the relevant gene is included (Figure 2). DECIPHER is a web platform developed to enable the annotation and sharing of anonymised phenotype-linked variants. It integrates essential genomics resources and provides visualisations and interactive tools to facilitate variant interpretation.

**Figure 2.**
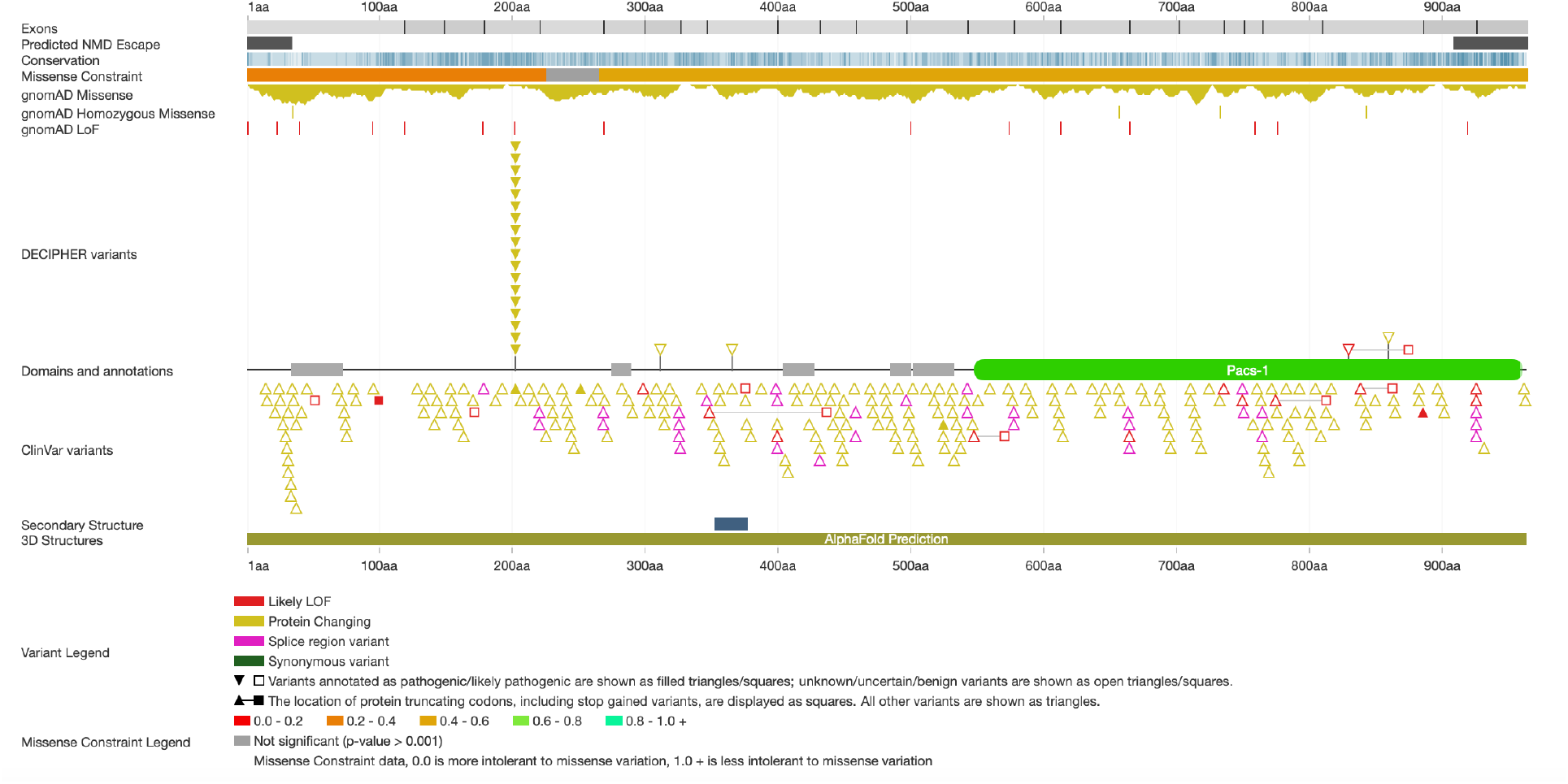
DECIPHER protein view for the gene PACS1 (13), particularly highlighting a single recurrent pathogenic variant. The web-based interface at www.deciphergenomics.org is interactive and regularly updated. Data is pulled from multiple bioinformatic resources. Note recurrent missense variant in ‘DECIPHER variants’ track, associated with disease in this gene (17).

The DECIPHER protein view for a given gene is an information dense graphical representation including, for example, exon boundaries, predicted NMD escape regions, conservation levels, missense constraint and protein domains. Annotation includes gnomAD (14) variants from global populations, as well as DECIPHER and ClinVar (15) variants from diagnostic testing (including reported pathogenicity).

Use of the DECIPHER protein view allows curators to easily put individual-level gene-disease information from publications into a wider context, encompassing molecular/gene-level data, disease-associated/diagnostic variation and population-level normal variation.

The function of the gene as defined by UniProt (16) is also shown in DECIPHER (13,16). This is used in the curation process to determine if the proposed gene-disease relationship is likely to be biologically plausible.

DECIPHER (13) is a constantly updated resource. The live website interface is often used in the curation process to interrogate the above data further. However, a snapshot of the protein view used at the time of curation is recorded for reference, to enable comparison with the most recent data at the time of re-curation.

#### Variant consequence per allele

For a given allelic requirement, SO terms for the variant consequence are used (11,12). This may be recorded as inferred or evidence based. Inferred includes, for example, when a variant is predicted to result in nonsense mediated decay.

Evidence based is usually used for biological functional studies such as demonstration of absent protein expression. Computational evidence may also be taken into account, such as modelling of protein structure. Terms for both altered protein (for protein-coding genes) or altered RNA level (for non-protein coding genes) are included. For example, a missense variant may be recorded as altered_gene_product structure (SO:0002318).

As SO terms are part of an ontology, higher level terms may be inferred for a recorded variant consequence. For example, absent_gene_product (SO:0002317) is a child of decreased_gene_product_level (SO:0002316), which itself is a child of altered_gene_product_level (SO:0002314).

#### Mechanism

Enabling a precise definition of the mechanism of disease to be captured is a crucial function of G2P. This follows the definitions laid out by Backwell & Marsh (18). The mechanism is initially recorded in broad categories depending on the protein level consequence of variants reported. These include ‘Loss of function’, ‘Dominant negative’ and ‘Gain of function’. There are also categories for ‘Undetermined’ and ‘Undetermined non-loss-of-function’, reflecting the fact that there are often cases where a mechanism is clearly not a loss of function, but where is difficult to distinguish between dominant-negative and gain-of-function effects from available evidence. It is recorded whether these are inferred or from functional evidence.

#### Synopsis of mechanism

As per Backwell & Marsh (18), the complex effects of disease-associated variants are not fully captured using broad terms such as ‘loss of function’ alone. Therefore, G2P records a more detailed synopsis of the inferred or evidence-based molecular mechanism. For example, ‘Destabilising loss of function’ or ‘Interaction-disrupting loss of function’. There is often insufficient evidence for newly described disorders to record this information. This may be completed when functional studies become available for a particular condition/variant(s).

The process of determining the likely mechanism for a given disorder is complex. Multiple lines of evidence are reviewed, where available. These may include, for example, observations of variant clustering and *in vitro/in vivo* functional assays. Tools predicting likely mechanism may be used for guiding assessments, although these should not be relied upon alone (19). MaveDB is interrogated for relevant multiplexed assays of variant effect (MAVEs) (20). Free text fields are currently used to record analysis of this evidence during the curation process. For example, it is critical to evaluate if a MAVE assay is relevant to the disorder being curated, especially whether the multiplexed assay truly reflects the mechanism of disease *in vivo*. Hence, we have prioritised curation of mechanism in G2P. Curation discussion may include information such as the functional domains assayed, how well it replicates the mechanism of disease for the stated gene-disease pair, and which tissue/cell-line is relevant.

### Disease entity

#### Clinical Phenotype

The reported clinical phenotype is recorded per publication. This includes the number of families/individuals reported, including information on consanguinity and/or ethnicity if relevant. G2P records phenotype data in the form of HPO terms (9), which are standardised and machine readable. However, it is also useful for curation purposes to record descriptive free text regarding the phenotype. For example, the proportion of individuals reported with a given phenotypic feature (i.e. variable expressivity), whether the phenotype is clinically distinctive and/or consistent, and if there is evidence for incomplete penetrance. The clinical phenotype is of crucial importance in determining the confidence level for a given gene-disease association.

#### Disease name

Genetic disease naming is a complex topic, in part reflecting the evolution in knowledge from clinical descriptions to molecular diagnosis. Conditions which have been well-defined clinically in the past may be known by an eponymous name such as Noonan syndrome (21). However, this naming system does not reflect the molecular basis of disease. This is especially true for conditions such as Noonan, where a similar phenotype is now known to result from variants in multiple genes.

The dyadic naming system suggested by Biesecker *et al*. (22) aims to address these issues by including a gene symbol with a phenotypic descriptor – for example, *PTPN11-*related Noonan syndrome. Ideally, a precise clinically relevant phenotypic descriptor is used. For example, *AMOTL1*-related orofacial clefting, cardiac anomalies, and tall stature.

G2P records disease names following this dyadic approach. However, the process of naming a condition is not straightforward. We recognise that an international collaborative approach is needed to address this topic, as other curation resources may define diseases differently, or use an alternate naming convention (7). If a disorder has been named – in a compatible format – by another group, G2P aims to use this name, to enhance standardisation across resources. In some cases, the gene symbol may be added to an existing disease name to maintain the dyadic structure. Mapping to other resources is added, where available, for example to OMIM (Online Mendelian Inheritance in Man) Morbid IDs and Mondo IDs (23,24). Disease synonyms from these and other sources are also included.

G2P is updated at least monthly with information from the latest research publications. Many newly defined conditions do not have a recognised disease name. In this case, the curation group agrees on a dyadic name reflecting the most pertinent phenotypic features and plans to submit these for Mondo accessioning to enable reuse.

#### Agreed confidence category

A confidence attribute is assigned to each G2P entry to indicate the likelihood that the gene-disease association is true. G2P now uses the gene-disease validity terms developed by GenCC (7). Gene-disease associations curated as ‘Definitive’, ‘Confirmed’ and ‘Moderate’ are used by several groups in clinical reporting, for example in the DDD study (3). Assertions with the confidence term ‘Limited’ are excluded from clinical reporting, however variants found in this group may be useful in the research setting and they may be promoted to a higher confidence level as further evidence becomes available. ‘Disputed’ and ‘Refuted’ are also used in G2P to indicate previously reported gene-disease links that should now be excluded from research or clinical use.

#### Panel

G2P is grouped into broad panels, which each focus on a disease grouping or defined category of clinical presentation of relevance to the clinical diagnosis of Mendelian disease. The curation structure outlined here is presently used by the DDG2P curation group, although it is anticipated other G2P panels will adopt it in future.

#### Evidence

Links to the original peer-reviewed publications analysed during the curation process are recorded, generally in the form of PubMed ID (25) and title for ease of reference. Manuscripts from non-peer reviewed sources such as MedRxiv are generally not included, except in exceptional circumstances.

## Discussion

The structured curation process described here uses LGMDE threads to define genetically-determined disorders from the molecular level through to clinical phenotype.

This creates a focus on disease mechanism and differentiates G2P from other initiatives recording gene-disease associations. For example, PanelApp (26) records an allelic requirement and phenotype terms in association with a gene to define a disease. The granularity of data in the G2P curation template should allow for more accurate interpretation of genomic results, especially where the disease mechanism is complex or multiple disorders are associated with a given gene. Coupling disease-mechanism and inheritance-based variant filtering can significantly reduce the noise associated with large, highly variable genes, by focussing interpretation time only on specific known variants that are consistent with the disease mechanism.

G2P covers a wide spectrum of genetic disease through a limited number of large domain-specific panels. Other sources, such as PanelApp (26), may define many more panels in association with more specific phenotypes/diseases. These often contain small numbers of genes, to decrease the number of variants of uncertain clinical significance returned. This requires curation regarding which panels are applied to which patients, as well as curation of the individual gene-disease associations. Furthermore, application of small panels is likely to have a higher specificity (fewer false positives) but lower sensitivity (fewer true positives) than larger panels. In contrast, G2P allows for scalable high-throughput filtering of large numbers of variants across more genes through precise disease definition. This is particularly relevant for disease domains where there is high locus and allelic heterogeneity, such as developmental disorders.

The curation process outlined here demonstrates a transparent, standardised and comprehensive method for collating and critically assessing data relevant to each G2P entry. We also include a more detailed analysis of disease mechanism, particularly building on the work of Backwell and Marsh (18). Moving from broad categorisations, such as loss of function, towards precise definitions of the molecular effects of disease-associated variants is increasingly important in the context of targeted therapeutics. These include gene therapy, targeting of antisense oligonucleotides and identification of other drug targets.

However, despite its importance, there is often insufficient evidence on first curation of a newly described disorder to precisely determine the disease mechanism. Relevant data fields need to be retrospectively completed when further functional studies become available. This emphasises the importance of regular review of disease entries, and re-curation with new evidence. The curation template used here, and the G2P database structure, allow for iterative re-curation in this manner.

The updated curation template described here is in active use and the online G2P database is now undergoing further development to reflect these changes. Future planned updates include the provision of data downloads both of gene records and full panel summaries, implementation of a web-based curation tool including standardised functional data, and incorporation of automated manuscript search and annotation.

## Conclusions

In summary, we present a structured process for the curation of gene-disease assertions, with precise definitions of the underlying molecular mechanism. This is compatible with international initiatives aimed at harmonising gene curation (7). The process defines rigorous, evidence-based gene-disease associations, which will prove useful in a range of applications, including clinical diagnostics and targeted therapeutics.

## Data Availability

The G2P data curation template is available as a supplementary file and on the G2P FTP site (http://ftp.ebi.ac.uk/pub/databases/gene2phenotype/).The datasets generated through the process described in the current study are available in the G2P repository, www.ebi.ac.uk/gene2phenotype/downloads.

http://ftp.ebi.ac.uk/pub/databases/gene2phenotype/

http://www.ebi.ac.uk/gene2phenotype/downloads

## Abbreviations

G2P: Gene2Phenotype
DDD: Deciphering Developmental Disorders
LGMDE: Locus-Genotype-Mechanism-Disease-Evidence
DDG2P: Developmental Disorders Gene2Phenotype
GenCC: Gene Curation Coalition
HGNC: HUGO Gene Nomenclature Committee
HPO: Human Phenotype Ontology
SO: Sequence Ontology
NMD: Nonsense Mediated Decay
OMIM: Online Mendelian Inheritance in Man

## Declarations

### Ethics approval and consent to participate

Not applicable

### Consent for publication

Not applicable

### Availability of data and materials

DECIPHER is found at https://deciphergenomics.org.

### Authors’ contributions

TMY, MA, LT, SEH, ECU, RJH, JAM, CFW and HVF made substantial contributions to the conception and design of the curation template. MA and LT iteratively drafted the curation template document. TMY drafted the initial manuscript. All authors read, revised and approved the final manuscript.

## Acknowledgements

We thank our collaborators who attended of the PARADIGM G2P Gene Curation Workshop and reviewed the curation template: James Ware, Professor of Cardiovascular and Genomic Medicine, Imperial College London, Ian Simpson, Professor of Biomedical Informatics & Director UKRI CDT in Biomedical Artificial Intelligence, University of Edinburgh, Andy Yates, Team Leader, Genomics Technology Infrastructure, EMBL-EBI, Sarah Wynn, Chief Executive Officer, Unique, Rare Chromosome Disorder Support Group, London, Adam Frankish, Manual Genome Annotation Coordinator, EMBL-EBI, Tariro Chatiza, PhD student, University of Edinburgh, Richard Chin, Professor of Paediatric Neurology and Clinical Epidemiology, Clinical Director of the Muir Maxwell Epilepsy Centre, University of Edinburgh, Diana Lemos, Senior Bioinformatics Developer, EMBL-EBI, Ola Austine, Bioinformatics Software Developer, EMBL-EBI, Nicky Whiffin, Associate Professor and Group Leader at the Big Data Institute and the Wellcome Centre for Human Genetics, University of Oxford, Alison Meynert, Senior Research Fellow, IGC Bioinformatics Analysis Core Manager, University of Edinburgh, Angharad Roberts, Clinical Senior Lecturer in Cardiovascular Genetics, Imperial College London, Jane Loveland, Annotation Project Leader, EMBL-EBI, Katharine Fitzpatrick, PARADIGM Project Manager, University of Exeter, Kat Josephs, Specialist registrar in Clinical Genetics and a Clinical Research Fellow in the Cardiovascular Genetics and Genomics group, Imperial College London, Massimo Mangino, Imperial College London, Julia Foreman, DECIPHER Project Leader, EMBL-EBI, We thank the clinical genomics community for ongoing curation suggestions. We thank Professor David FitzPatrick for initially setting up and further developing G2P.

