## Supplementary Materials for "Curating genomic disease-gene relationships with Gene2Phenotype"

### G2P CURATION RECORD VERSION 7.2

| |
|---|
| **Version:** |
| **Date created:** |
| **Date modified:** |

**Gene symbol (HGNC)**

**Gene-disease name (dyadic naming system)**

**List of associated PMID and titles**

**Protein function as described by UniProt**
**(copy and paste from DECIPHER Overview)**

**Gene currently in G2P https://www.ebi.ac.uk/gene2phenotype?**

Yes ☐ If yes, what category and which panel(s)?

No ☐ Please specify which panel(s)

**Gene currently in GenCC https://search.thegencc.org/?**

Yes ☐ If yes, what category or categories?

No ☐

**Number of families and associated phenotype per PMID (include information on consanguinity, specific ethnicity, if relevant)**

| PMID | No. of families | Notes |
|---|---|---|

**Allelic requirement**

| | | |
|---|---|---|
| Autosomal Dominant | monoallelic_autosomal | ☐ |
| Autosomal Recessive | biallelic_autosomal | ☐ |
| X-linked | monoallelic_X_heterozygous | ☐ |
| X-linked | monoallelic_X_hemizygous | ☐ |
| Y-linked | monoallelic_Y_hemizygous | ☐ |
| Mitochondrial | mitochondrial | ☐ |
| PAR dominant | monoallelic_PAR | ☐ |
| PAR recessive | biallelic_PAR | ☐ |

G2P CURATION RECORD VERSION 7.2

#### Clinical Phenotype

| |
|---|
| *Summary of the reported clinical phenotype (include information on variable penetrance)* |

#### Cross cutting modifier

| | |
|---|---|
| Typically de novo | ☐ |
| Typically mosaic | ☐ |
| Typified by incomplete penetrance | ☐ |
| Imprinted region | ☐ |
| Potential secondary finding (including ACMG Secondary Findings and/or late onset conditions) | ☐ |
| Displays anticipation | ☐ |
| Restricted variant set | ☐ |

#### Types of variants reported

| **Frameshift & nonsense variants** | **Comment on NMD triggering/escaping if necessary** | **De novo** | **Inherited** | **Unknown inheritance** |
|---|---|---|---|---|
| frameshift_variant | | ☐ | ☐ | ☐ |
| stop_gained | | ☐ | ☐ | ☐ |
| **Splice variants** | **Comment on NMD triggering/escaping if necessary** | **De novo** | **Inherited** | **Unknown inheritance** |
| splice_region_variant | | ☐ | ☐ | ☐ |
| splice_acceptor_variant | | ☐ | ☐ | ☐ |
| splice_donor_variant | | ☐ | ☐ | ☐ |
| **Missense & inframe variants** | **Comment on domain/region** | **De novo** | **Inherited** | **Unknown inheritance** |
| missense_variant | | ☐ | ☐ | ☐ |
| inframe_insertion | | ☐ | ☐ | ☐ |
| inframe_deletion | | ☐ | ☐ | ☐ |
| **Other variants** | | **De novo** | **Inherited** | **Unknown inheritance** |
| start_lost | | ☐ | ☐ | ☐ |
| intergenic_variant | | ☐ | ☐ | ☐ |
| intron_variant | | ☐ | ☐ | ☐ |
| synonymous_variant | | ☐ | ☐ | ☐ |
| Stop lost | | ☐ | ☐ | ☐ |
| Whole/partial gene deletion | | ☐ | ☐ | ☐ |
| Whole/partial gene duplication | | ☐ | ☐ | ☐ |
| short_tandem_repeat_change | | ☐ | ☐ | ☐ |
| ncRNA | | ☐ | ☐ | ☐ |
| **Variants in regulatory regions** | | **De novo** | **Inherited** | **Unknown inheritance** |
| 5_prime_UTR_variant | | ☐ | ☐ | ☐ |
| 3_prime_UTR_variant | | ☐ | ☐ | ☐ |
| regulatory_region_variant | | ☐ | ☐ | ☐ |

#### DECIPHER Protein View

(logged-out version)

#### Variant consequence per allele for relevant allelic requirement (from above)

**Hierarchy of SO disease-associated variant consequence terms (described in https://www.ncbi.nlm.nih.gov/pmc/articles/PMC10104222/pdf/nihpp-2023.03.30.23287948v1.pdf)**

| ***Altered protein for protein-coding genes or altered RNA level for non-protein coding genes*** | **Inferred** | **Evidence** |
|---|---|---|
| Altered_gene_product_level SO:0002314 | ☐ | ☐ |
| • Decreased_gene_product_level SO:0002316 | ☐ | ☐ |
| - Absent_gene_product SO:0002317 | ☐ | ☐ |
| • Increased_gene_product_level SO:0002315 | ☐ | ☐ |
| Altered_gene_product_structure SO:0002318 | ☐ | ☐ |
| Altered gene product function (eg. missense variants or frameshift and nonsense variants and in-frame indels escaping NMD, UTR variants changing start site) | ☐ | ☐ |
| Uncertain | ☐ | ☐ |

#### Mechanism

| | **Description** | **Inferred** | **Evidence** |
|---|---|---|---|
| Loss of function | *Loss-of-function variants involve a loss of the normal biological function of a protein. Often these are nonsense or frameshift mutations that introduce premature stop codons. Due to nonsense-mediated decay of the resulting mRNAs, most premature stop codons will result in no protein being produced, rather than a truncated protein. However, there are also many examples of loss-of-function variants that change the amino acid sequence and result in non-functional protein products. These mutations can cause a complete loss of function (**amorphic**), analogous to a protein null mutation, or only a partial loss of function (**hypomorphic**). May also include variants in regulatory regions.* | ☐ | ☐ |
| Dominant negative | *Dominant-negative variants involve the mutant protein directly or indirectly blocking the normal biological function of the wild-type protein (**antimorphic**). They can thus cause a disproportionate (>50%) loss of function, even though only half of the protein is mutated eg. heterozygous variants in COL1A1 that disrupt the triple collagen helix.* | ☐ | ☐ |
| Gain of function | *Gain-of-function variants have their phenotypic effect because the mutant protein does something different than the wild-type protein. Often, these variants cause disease by increasing protein activity (**hypermorphic**) or introducing a completely new function (**neomorphic**), but the specific molecular mechanisms underlying gain-of-function mutations can be complex. May also include variants in regulatory regions.* | ☐ | ☐ |
| Undetermined non-loss-of-function | *Very often it is difficult to distinguish between dominant negative and gain of function, but it is clearly a non-loss-of-function mechanism (e.g. from co-expression experiments showing a damaging effect from the mutant allele).* | ☐ | ☐ |
| Undetermined | | ☐ | ☐ |

#### Categorisation of mechanism

*Refer to Backwell & Marsh, 2022 (PMID 35395171) for help and examples)*
*If possible, categorise into:*

| | Inferred | Evidence |
|---|---|---|
| Destabilising LOF | ☐ | ☐ |
| Interaction-disrupting LOF | ☐ | ☐ |
| Loss of activity LOF (*e.g.* active site mutation) | ☐ | ☐ |
| LOF due to protein mislocalisation | ☐ | ☐ |
| Assembly-mediated dominant negative (*i.e.* poisoning via mutant subunit) | ☐ | ☐ |
| Competitive dominant-negative | ☐ | ☐ |
| Assembly-mediated GOF (*e.g.* channel activation via mutant subunit) | ☐ | ☐ |
| Protein aggregation (usually toxic GOF) | ☐ | ☐ |
| Local LOF (separation of function) leading to overall GOF (*e.g.* DNMT3A) | ☐ | ☐ |
| Other GOF (*e.g.* strengthening or gain of interactions, change in specificity, gain of post-translational modifications) | ☐ | ☐ |

*Provide details on altered variant consequence and mechanism classification, including references. Inferred consequence could also include predicted mechanism (e.g. Badonyi & Marsh, https://doi.org/10.1101/2023.09.08.556798), observations of mutation clustering*

#### Functional analysis of the variants

*Is there a MAVE or scalable functional assay for the gene? If so, what functional domains does it assay, how well does it replicate the mechanism of disease for the stated gene-disease pair, and what tissue/cell-line is it relevant to*

*Consider functional assays (in vitro, in vivo), animal models, patient cell lines*

#### Additional comments

*Consider previous publications, mutational landscape on DECIPHER and gnomAD*

#### Discussion of current gene-disease name and additional information, if applicable

#### Synonyms:

| OMIM number (gene) | MONDO number |
|---|---|

#### Panel

| | |
|---|---|
| Cancer | ☐ |
| DDG2P | ☐ |
| Eye | ☐ |
| Neonatal | ☐ |
| Obesity | ☐ |
| PNG2P | ☐ |
| Skeletal | ☐ |
| Skin | ☐ |

#### Agreed confidence category

| | |
|---|---|
| Definitive | ☐ |
| Strong | ☐ |
| Moderate | ☐ |
| Limited | ☐ |
| Disputed | ☐ |
| Refuted | ☐ |

#### Changed from (if relevant)

| | |
|---|---|
| Definitive | ☐ |
| Strong | ☐ |
| Moderate | ☐ |
| Limited | ☐ |

**Description of G2P confidence categories**

**Definitive**: The role of this gene in this particular disease has been repeatedly demonstrated in both the research and clinical diagnostic settings, and has been upheld over time (at least 2 independent publication over 3 years' time). No convincing evidence has emerged that contradicts the role of the gene in the specified disease. (previously labelled as confirmed).

**Strong**: The role of this gene as a monogenic cause of disease has been repeatedly and independently demonstrated providing very strong convincing evidence in humans and no conflicting evidence for this gene's role in this disease. (previously labelled as probable)

**Moderate**: There is moderate evidence in humans to support a casual role for this gene in this disease with no contradictory evidence. The body of evidence is not large (e.g possibly only one key paper) but appears convincing enough that the gene-disease pair is likely to be validated with additional evidence in the near future.

**Limited**: Little human evidence exists to support a casual role for this gene in this disease, but not all evidence has been refuted. For example, there may be a collection of rare missense variants in humans but without convincing functional impact, segregration data that could either arise by chance (e.g across one or two meioses) or does not implicate a single gene, or functional data without direct recapitulation of the phenotype. Overall, the body of evidence does not meet contemporary criteria for claiming a valid association with disease. The majority are probably false associations. (previously labelled as possible).

Inheritance modifiers are described in detail by Roberts et al https://doi.org/10.1016/j.gim.2023.101029.
